# Quantification of occupational and community risk factors for SARS-CoV-2 seropositivity among healthcare workers in a large U.S. healthcare system

**DOI:** 10.1101/2020.10.30.20222877

**Authors:** Julia M. Baker, Kristin N. Nelson, Elizabeth Overton, Benjamin A. Lopman, Timothy L. Lash, Mark Photakis, Jesse T. Jacob, John Roback, Scott K. Fridkin, James P. Steinberg

## Abstract

**Background:** Quantifying occupational risk factors for SARS-CoV-2 infection among healthcare workers can inform efforts to improve healthcare worker and patient safety and reduce transmission. This study aimed to quantify demographic, occupational, and community risk factors for SARS-CoV-2 seropositivity among healthcare workers in a large metropolitan healthcare system.

**Methods:** We analyzed data from a cross-sectional survey conducted from April through June of 2020 linking risk factors for occupational and community exposure to COVID-19 with SARS-CoV-2 seropositivity. A multivariable logistic regression model was fit to quantify risk factors for infection. Participants were employees and medical staff members who elected to participate in SARS-CoV-2 serology testing offered to all healthcare workers as part of a quality initiative, and who completed a survey on exposure to COVID-19 and use of personal protective equipment. Exposures of interest included known demographic risk factors for COVID-19, residential zip code incidence of COVID-19, occupational exposure to PCR test-positive healthcare workers or patients, and use of personal protective equipment. The primary outcome of interest was SARS-CoV-2 seropositivity.

**Results:** SARS-CoV-2 seropositivity was estimated to be 5.7% (95% CI: 5.2%-6.1%) among 10,275 healthcare workers. Community contact with a person known or suspected to have COVID-19 (aOR=1.9, 95% CI:1.4-2.5) and zip code level COVID-19 incidence (aOR: 1.4, 95% CI: 1.0-2.0) increased the odds of infection. Black individuals were at high risk (aOR=2.0, 95% CI:1.6-2.4). Overall, occupational risk factors accounted for 27% (95% CI: 25%-30%) of the risk among healthcare workers and included contact with a PCR test-positive healthcare worker (aOR=1.2, 95% CI:1.0-1.6).

**Conclusions:** Community risk factors, including contact with a COVID-19 positive individual and residential COVID-19 incidence, are more strongly associated with SARS-CoV-2 seropositivity among healthcare workers than exposure in the workplace.

## Introduction

Healthcare workers (HCWs) are presumed to be at high risk for coronavirus disease 2019 (COVID-19) through occupational exposure to infected patients or coworkers. Studies have reported a wide range of seroprevalence of SARS-CoV-2, the virus that causes COVID-19, among HCWs. This variation has in part been attributed to differential risk of exposure related to COVID-19 incidence in the community.^1^ Indeed, recent studies have shown that a substantial number of infections among HCWs could not be traced to occupational exposures ^2^ and that community exposures were as or more strongly associated with infection.^3^ While previous studies have compared seroprevalence in HCWs with that of the general population, few have rigorously considered workplace risk factors alongside community risk factors among HCWs to estimate their relative contribution to overall infection risk.^4,5^

Accounting for the role of community risk, which may be large, is especially important because reports of occupational risk factors for SARS-CoV-2 infection among HCWs have been inconsistent.^3-8^ While some studies have shown weak associations between involvement in clinical care, care of COVID-19 patients, and exposure to coworkers with COVID-19, ^6-8^ others have shown that these are, in fact, risk factors for infection.^4,5^ Moreover, previous studies have not accounted for potential participation bias, though have cited it as a major limitation.^1,2,4,7^ Adjusting for participation bias while considering both workplace and community risk factors for infection can bring us closer to an accurate understanding of which workplace exposures confer the highest risk of infection for HCWs. This information can inform strategies to protect HCWs as the COVID-19 pandemic continues.

We aimed to quantify occupational, community, and demographic risk factors for SARS-CoV-2 seropositivity among HCWs in a large university-based healthcare system, adjusting for possible bias due to voluntary participation in testing. We estimate the proportion of risk attributable to occupational factors among HCWs.

## Methods

We analyzed data from Emory Healthcare, which includes 11 hospitals, 250 provider locations, and approximately 25,000 employees and medical staff members based in the Atlanta, Georgia, metropolitan area.

A voluntary serological survey was conducted among employees and medical staff members from April 19 through June 26, 2020 to inform process improvement activities. At the time of testing, HCWs completed a survey describing use of personal protective equipment (PPE) and possible exposure to COVID-19 inside and outside the workplace. These data were combined with employee demographic data. To assess community exposure to COVID-19, we used data from the Georgia Department of Public Health to calculate COVID-19 cumulative incidence by zip code and week. As a measure of exposure in their community, we assigned each participant the COVID-19 cumulative incidence in their zip code of residence two weeks prior to their test date to account for the lag from the time of infection to seroconversion (eFigure 1 in Supplement). The serologic test used to analyze participant samples was developed at the Emory Medical Laboratory and measures IgG antibody to the receptor binding domain of the SARS-CoV-2 spike protein.^9^

We fit a logistic regression model to estimate adjusted odds ratios (aORs) between potential risk factors and SARS-CoV-2 seropositivity. Model predictors (eTable 1 in Supplement) included demographics (age group; race; ethnicity), community exposure (cumulative incidence of COVID-19 by residential zip code two weeks before testing date; contact with confirmed/suspected COVID-19 cases outside the workplace), and occupational factors (workplace role and location; contact with COVID-positive patients and staff; use of PPE). Workplace location was self-reported and described the location(s) where a HCW spent the most time during the COVID-19 pandemic. Since over one-third (36.4%) of HCWs reported spending equal time in more than one location, the workplace location variable was structured as a hierarchy. We ranked locations based on the anticipated risk of encounters with COVID-19 positive patients (see eTable 1 in Supplement for details) with high encounter risk locations, such as the emergency department and COVID-19 focused units, highest and no patient contact and working from home ranked lowest. HCWs were categorized into the highest-ranking location where they reported working.

To account for potential selective participation, we used inverse probability of participation weighting to assess whether our results were sensitive to differences between those tested and all employees by age, race, and sex. We used the joint distribution of the age, race, and sex of all employees to weight individuals in the regression analysis such that demographic groups who were over-represented among survey participants compared to all employees were down-weighted, and vice versa. We calculated aORs using the weighted data and 95% confidence intervals (CIs) accounting for the weights.

Lastly, the fraction of infection attributable to occupational risk factors was estimated using a model-based approach to calculating the population attributable fraction for the combined set of all occupational risk factors.^10^ The 95% CIs for the attributable fraction was estimated by applying a normal distribution to the predicted probability of seropositivity for each survey participant and generating 1,000 bootstrap samples from this distribution. From these samples, we calculated the resulting distribution of the attributable fraction to calculate a 95% CI around the median attributable fraction. Analyses were conducted in R version 4.0.2 using the *survey, boot* and *zipcode* packages. The code for this analysis is available at https://github.com/lopmanlab/emory-hcw-serosurvey.

This study was approved by the Emory University Institutional Review Board.

## Results

Among 10,275 participating HCWs (35% of the workforce), the SARS-CoV-2 seroprevalence was 5.7% (95%CI: 5.2%-6.1%). A total of 665 participants were missing zip code data, leaving 9,610 participants available for further analysis.

The majority (71%) of participants were tested in May 2020 (eTable 2 in Supplement). Over three-quarters of participants were female (77.6%) with female and male participants having similar seropositivity (5.7% and 5.9%, respectively). Black HCW were notably underrepresented in the serosurvey, comprising 30% of those who volunteered for antibody testing and 49% of the workforce. Prior confirmed COVID-19 infection was reported for 133 participants (1.4%). Forty-four percent (245/555) of seropositive HCWs reported no fever or COVID-19-like symptoms since February 1, 2020.

Contact with a person known or suspected to have COVID-19 outside the workplace (aOR=1.9, 95% CI:1.4-2.5) and race were most strongly associated with seropositivity (Table 1). Black (aOR=2.0, 95%CI:1.6-2.4) and multiracial (aOR:1.7, 95%CI:0.8-3.4) HCWs had higher odds of infection than white HCWs. Higher residential zip code incidence of COVID-19 infection (aOR=1.4, 95%CI:1.0-2.0) was also associated with seropositivity. Age under 30 years was associated with seropositivity (aOR 1.3, 95% CI 0.9, 1.8).

**Table 1.** Association of demographic, community, and occupational risk factors with seropositivity in unadjusted and adjusted analyses (n=9,610).

| Factor | Total sample<br>N (%) | Sero-<br>positive<br>N (%) | Unadjusted<br>OR (95% CI) | Adjusted<br>(unweighted)<sup>a</sup><br>OR (95% CI) | Adjusted<br>(weighted, to<br>adjust for<br>participation)<sup>b</sup><br>OR (95% CI) |
| --- | --- | --- | --- | --- | --- |
| **Demographic and community factors** |  |  |  |  |  |
| Age group |  |  |  |  |  |
| 60+ | 1153 (12.0) | 57 (4.9) | 1.0 (ref) | 1.0 (ref) | 1.0 (ref) |
| 50-59 | 1805 (18.8) | 93 (5.2) | 1.0 (0.7, 1.5) | 0.9 (0.7, 1.3) | 1.0 (0.7, 1.4) |
| 40-49 | 2118 (22.0) | 125 (5.9) | 1.2 (0.9, 1.7) | 1.1 (0.8, 1.5) | 1.0 (0.7, 1.4) |
| 30-39 | 2953 (30.7) | 171 (5.8) | 1.2 (0.9, 1.6) | 1.1 (0.8, 1.6) | 1.1 (0.8, 1.5) |
| <30 | 1581 (16.5) | 109 (6.9) | 1.4 (1.0, 2.0) | 1.4 (1.0, 2.0) | 1.3 (0.9, 1.8) |
| Race |  |  |  |  |  |
| White | 5263 (54.8) | 226 (4.3) | 1.0 (ref) | 1.0 (ref) | 1.0 (ref) |
| Black | 2860 (29.8) | 238 (8.3) | 2.0 (1.7, 2.4) | 2.0 (1.6, 2.5) | 2.0 (1.6, 2.4) |
| Asian | 1133 (11.8) | 60 (5.3) | 1.2 (0.9, 1.7) | 1.2 (0.9, 1.6) | 1.1 (0.8, 1.5) |
| Multiracial | 132 (1.4) | 10 (7.6) | 1.8 (0.9, 3.4) | 1.8 (0.9, 3.3) | 1.7 (0.8, 3.4) |
| American Indian/Alaska Native | 33 (0.3) | --<sup>d</sup> | 1.4 (0.2, 4.8) | 1.6 (0.3, 5.5) | 1.1 (0.3, 4.9) |
| Native Hawaiian/Pacific Islander | 16 (0.2) | --<sup>d</sup> | --<sup>e</sup> | --<sup>e</sup> | --<sup>e</sup> |
| Not specified | 173 (1.8) | 19 (11.0) | 2.7 (1.6, 4.4) | 2.8 (1.6, 4.7) | 3.0 (1.8, 5.2) |
| Ethnicity |  |  |  |  |  |
| Non-Hispanic | 9206 (95.8) | 534 (5.8) | 1.0 (ref) | 1.0 (ref) | 1.0 (ref) |
| Hispanic | 404 (4.2) | 21 (5.2) | 0.9 (0.6, 1.4) | 0.8 (0.5, 1.2) | 0.7 (0.4, 1.1) |
| Community contact with<br>confirmed/suspected positive<br>individual(s) |  |  |  |  |  |
| No | 8862 (92.2) | 478 (5.4) | 1.0 (ref) | 1.0 (ref) | 1.0 (ref) |
| Yes | 748 (7.8) | 77 (10.3) | 2.0 (1.6, 2.6) | 1.7 (1.3, 2.2) | 1.9 (1.4, 2.5) |
| Residential COVID-19 incidence<sup>c</sup> | 1.70 (mean) | 1.72 (mean) | 1.4 (1.0, 1.9) | 1.2 (0.9, 1.7) | 1.4 (1.0, 2.0) |
| **Occupational factors** |  |  |  |  |  |
| Caring for COVID-19 positive<br>patient(s) |  |  |  |  |  |
| No | 6049 (62.9) | 332 (5.5) | 1.0 (ref) | 1.0 (ref) | 1.0 (ref) |
| Yes | 3561 (37.1) | 223 (6.3) | 1.2 (1.0, 1.4) | 0.9 (0.7, 1.2) | 0.9 (0.7, 1.3) |
| Caring for patient(s) found to be<br>COVID-19 positive while not on<br>isolation precautions |  |  |  |  |  |
| No | 7893 (82.1) | 437 (5.5) | 1.0 (ref) | 1.0 (ref) | 1.0 (ref) |
| Yes | 1717 (17.9) | 118 (6.9) | 1.3 (1.0, 1.5) | 1.1 (0.8, 1.4) | 1.1 (0.8, 1.5) |
| Contact with other healthcare<br>worker found to be COVID-19<br>positive |  |  |  |  |  |
| No | 7878 (82.0) | 421 (5.3) | 1.0 (ref) | 1.0 (ref) | 1.0 (ref) |
| Yes | 1732 (18.0) | 134 (7.7) | 1.5 (1.2, 1.8) | 1.4 (1.1, 1.7) | 1.2 (1.0, 1.6) |

**Table 1. Association of demographic, community, and occupational risk factors with seropositivity in unadjusted and adjusted analyses (n=9,610) (continued).**
| Factor | Total sample N. (%) | Sero-positive N. (%) | Unadjusted OR (95% CI) | Adjusted (unweighted)<sup>a</sup> OR (95% CI) | Adjusted (weighted, to adjust for participation)<sup>b</sup> OR (95% CI) |
| --- | --- | --- | --- | --- | --- |
| Workplace location |  |  |  |  |  |
| No patient contact | 941 (9.8) | 43 (4.6) | 1.0 (ref) | 1.0 (ref) | 1.0 (ref) |
| Work from home | 267 (2.8) | 13 (4.9) | 1.1 (0.5, 2.0) | 1.0 (0.5, 1.9) | 1.0 (0.5, 1.9) |
| Other hospital area | 926 (9.6) | 52 (5.6) | 1.2 (0.8, 1.9) | 1.2 (0.8, 1.8) | 1.2 (0.8, 1.9) |
| OR/procedure | 672 (7.0) | 38 (5.7) | 1.3 (0.8, 2.0) | 1.2 (0.7, 1.9) | 1.0 (0.6, 1.7) |
| Outpatient clinical | 1675 (17.4) | 90 (5.4) | 1.2 (0.8, 1.7) | 1.2 (0.8, 1.8) | 1.3 (0.8, 2.0) |
| Inpatient not COVID-19 focused | 1594 (16.6) | 93 (5.8) | 1.3 (0.9, 1.9) | 1.4 (0.9, 2.1) | 1.3 (0.8, 2.1) |
| COVID-19 focused | 1750 (18.2) | 117 (6.7) | 1.5 (1.1, 2.2) | 1.5 (1.0, 2.2) | 1.5 (0.9, 2.4) |
| Emergency department | 1007 (10.5) | 67 (6.7) | 1.5 (1.0, 2.2) | 1.4 (0.9, 2.2) | 1.4 (0.9, 2.4) |
| Not specified | 778 (8.1) | 42 (5.4) | 1.2 (0.8, 1.8) | 1.2 (0.8, 1.9) | 1.1 (0.7, 1.9) |
| Workplace role |  |  |  |  |  |
| Other with no patient contact | 1812 (18.9) | 100 (5.5) | 1.0 (ref) | 1.0 (ref) | 1.0 (ref) |
| Nurse | 2976 (31.0) | 177 (5.9) | 1.1 (0.8, 1.4) | 1.0 (0.7, 1.3) | 1.0 (0.7, 1.4) |
| Physician | 1753 (18.2) | 87 (5.0) | 0.9 (0.7, 1.2) | 1.0 (0.7, 1.4) | 1.1 (0.7, 1.6) |
| Other direct care provider | 1423 (14.8) | 88 (6.2) | 1.1 (0.8, 1.5) | 1.0 (0.7, 1.4) | 1.0 (0.7, 1.4) |
| Advanced practice provider | 698 (7.3) | 36 (5.2) | 0.9 (0.6, 1.4) | 0.9 (0.6, 1.4) | 0.9 (0.5, 1.4) |
| Nurse technician | 346 (3.6) | 28 (8.1) | 1.5 (1.0, 2.3) | 0.9 (0.6, 1.5) | 0.9 (0.6, 1.6) |
| Radiology technician | 302 (3.1) | 21 (7.0) | 1.3 (0.8, 2.0) | 1.1 (0.7, 1.9) | 1.2 (0.7, 2.2) |
| Respiratory therapist | 114 (1.2) | --<sup>d</sup> | 1.0 (0.4, 2.0) | 0.7 (0.3, 1.6) | 0.9 (0.3, 2.4) |
| Environmental services | 35 (0.4) | --<sup>d</sup> | 1.6 (0.4, 4.6) | 1.1 (0.3, 3.3) | 0.9 (0.3, 3.2) |
| Not specified | 151 (1.6) | --<sup>d</sup> | 1.1 (0.5, 2.1) | 0.9 (0.4, 1.8) | 1.0 (0.5, 2.3) |
| Personal protective equipment use |  |  |  |  |  |
| As recommended | 6320 (65.8) | 370 (5.9) | 1.0 (ref) | 1.0 (ref) | 1.0 (ref) |
| Not as recommended | 221 (2.3) | 14 (6.3) | 1.1 (0.6, 1.8) | 0.9 (0.5, 1.6) | 1.1 (0.6, 2.0) |
| Unsure | 546 (5.7) | 22 (4.0) | 0.7 (0.4, 1.0) | 0.6 (0.4, 1.0) | 0.7 (0.4, 1.2) |
| Not specified | 2523 (26.3) | 149 (5.9) | 1.0 (0.8, 1.2) | 1.1 (0.9, 1.4) | 1.1 (0.9, 1.5) |

<sup>a</sup> Adjusted for all other factors shown in table.
<sup>b</sup> Adjusted for all other factors shown in table and accounting for participation bias using inverse probability of participation weighting for age group, race, and sex.
<sup>c</sup> The base-10 logarithm of cumulative COVID-19 incidence in a participant's zip code two weeks prior to the date they were tested.
OR: odds ratio; CI: confidence interval, ref: reference group
<sup>d</sup> Data suppressed because N < 10
<sup>e</sup> Odds ratio suppressed due to small sample size

In the workplace, participants who reported close contact with a COVID-positive HCW had increased odds of seropositivity (aOR=1.2, 95% CI:1.0-1.6). Although HCWs who reported caring for patients with COVID-19 did not have increased odds of seropositivity (aOR=0.9, 95% CI: 0.7, 1.3), working in clinical locations, such as inpatient non-COVID-19 focused areas (aOR=1.3, 95% CI: 0.8, 2.1), the emergency department (aOR=1.4, 95% CI: 0.9, 2.4) or COVID-focused units (aOR=1.5, 95% CI: 0.9, 2.4), was associated with higher odds of seropositivity. Respiratory therapists (aOR=0.9, 95% CI: 0.3, 2.4) and those who work in the operating room or procedure areas (aOR=1.0, 95% CI: 0.6, 1.7) were not at increased risk of being seropositive. Differences in seropositivity based on HCW role were generally present in the unadjusted but not the adjusted models. Accounting for participation bias did not result in appreciably different estimated associations (Table 1).

Overall, 27% (95% CI: 25%-30%) of risk could be attributed to the occupational risk factors included in our analysis.

## Discussion

In this large serological testing effort of U.S. HCWs in a university-based healthcare system, we found an overall SARS-CoV-2 seroprevalence of 5.7% (95% CI: 5.2%-6.1%) following the initial surge of the epidemic. This rate is similar to the overall 6.0% seroprevalence reported from 13 U.S. academic medical centers and is generally consistent with other estimates among HCWs in the early stages of the pandemic. ^1,2,7,11^ The percent of seropositive HCWs who reported no past COVID-19-like illness was also similar to past estimates of asymptomatic SARS-CoV-2 infections.^1^

We found that community and demographic factors--contact with a confirmed or suspected COVID-positive case, Black race, and age under 30 years--were more important predictors of seropositivity than occupational factors. Notably, racial disparities, now well-documented in the general population, ^12-14^ persist in HCWs ^1,15^ after accounting for other risk factors, underscoring the fundamental racial inequities that have become the hallmark of the COVID-19 pandemic. We partially adjusted for community risk by including zip code-level COVID-19 incidence in our model, but we are unable to account for more proximal factors likely to have contributed to higher risk of infection among Black HCWs, including higher likelihood of exposure at home or use of public transportation. Ultimately, these risk factors are tied to entrenched, systemic social processes that underlie many individual and population health disparities. ^16,17^

We found few strong risk factors for infection in the workplace. Those who reported caring for patients with COVID-19 infection and those working in procedure areas where aerosol generating procedures are routine were not more likely to be seropositive, supporting the efficacy of PPE practices for known COVID-19 patients. Working in clinical areas was associated with increased odds of seropositivity, although the aORs were imprecise. We were not able to determine if risk associated with workplace location was from patient exposure, including unsuspected COVID-19 infected patients or from co-workers. Risk from contact with HCWs later found to be COVID-19-positive could reflect transmission during pre-symptomatic or asymptomatic infection prior to universal use of masks or transmission in settings where masks were not worn. In our hospital system, contact tracing identified staff eating together in break rooms as a risk factor for transmission.

While there appeared to be a difference in seropositivity based on workplace role in the unadjusted model, aORs were similar for all workplace roles in the adjusted model, underscoring the importance of considering demographic factors in assessing seroprevalence risk. Other studies that have not controlled for demographic or other risk factors outside of the workplace have reported different seroprevalence rates depending on job role ^4,5^; these results need to be interpreted with caution. Overall, we found that 27% (95% CI: 25%-30%) of risk could be attributed to the set of occupational risk factors we considered (Table 1). While the majority of risk may originate from the community, workplace risk cannot be ignored. In this cross-sectional analysis including time periods before and after universal masking, we were not able to assess the impact of infection prevention policies in reducing infection risk among HCWs. Going forward, studies should investigate the role of specific exposures contributing to infection risk, including risk from HCW to HCW exposure, and the efficacy of interventions to prevent transmission. It is critical to know if the current level of interventions, including screening for asymptomatic viral shedding, PPE practices and efforts to prevent HCW to HCW transmission, have substantially reduced or eliminated the workplace risk identified in this and other studies.

There are several limitations to this analysis. First, serological testing was voluntary, which may introduce bias if groups more likely to participate were also more (or less) likely to be seropositive. We partially adjusted for participation using demographic characteristics of HCWs overall. This adjustment at least partially accounts for poor representation of Black HCWs, in whom seroprevalence was higher than the population average, among those who volunteered for testing. However, other factors related to infection risk may have influenced participation. Second, a test with imperfect specificity in a population where seroprevalence is low will likely result in some false positives which may bias aORs towards the null. Third, we could not account for rapidly evolving infection prevention practices early in the pandemic and social behavior inside or outside the workplace. Lastly, the large influx of COVID-19 patients caused major shifts in care delivery and personnel deployment. Many HCWs worked in multiple locations and even different roles.

In conclusion, using a model incorporating demographic, community, and occupational risk factors for infection, we quantified community and occupational risk of SARS-CoV-2 seropositivity in HCWs. We found that most of this risk is due to community exposure; ongoing efforts to keep the healthcare workforce safe should emphasize risk mitigation in and outside the workplace. After adjusting for a number of community and occupational risk factors, race remains a critical marker of infection risk. Future seroprevalence studies of HCWs need to account for these community and demographic factors.

## Data Availability

The data that support the findings of this study are not publicly available due to participant privacy. Requests for data should be submitted to the authors and require approval from Emory Healthcare.

## Acknowledgements

The authors are grateful to the Emory Healthcare employees and medical staff who volunteered for serology testing and completed the accompanying survey. We thank Laura Edison and Melissa Tobin-D’Angelo from the Georgia Department of Public Health for providing zip code level data on COVID-19 incidence and for their review of the manuscript. The authors have no conflicts of interest to disclose.

## Funding statement

Kristin N. Nelson and Benjamin A. Lopman received funding support from the Emory COVID-19 Response Collaborative. Julia M. Baker received funding support from the National Institute of Allergy and Infectious Diseases (T32AI074492).

## Notes

### Competing Interest Statement

The authors have declared no competing interest.

### Author Declarations

This study was approved by the Emory University Institutional Review Board.

