## Supplemental Material for "Quantification of occupational and community risk factors for SARS-CoV-2 seropositivity among healthcare workers in a large U.S. healthcare system"


### Supplement

**eFigure 1.** Number of study participants (A) and cumulative incidence of COVID-19 (B) by zip code [a]

**eTable 1.** Details of data sources and factor descriptions

**eTable 2.** Additional descriptive statistics for survey participants. (n=9,610)

**eTable 3.** STROBE statement- checklist of items that should be included in reports of observational studies

**eFigure 1. Number of study participants (A) and cumulative incidence of COVID-19 (B) by zip code [a]**

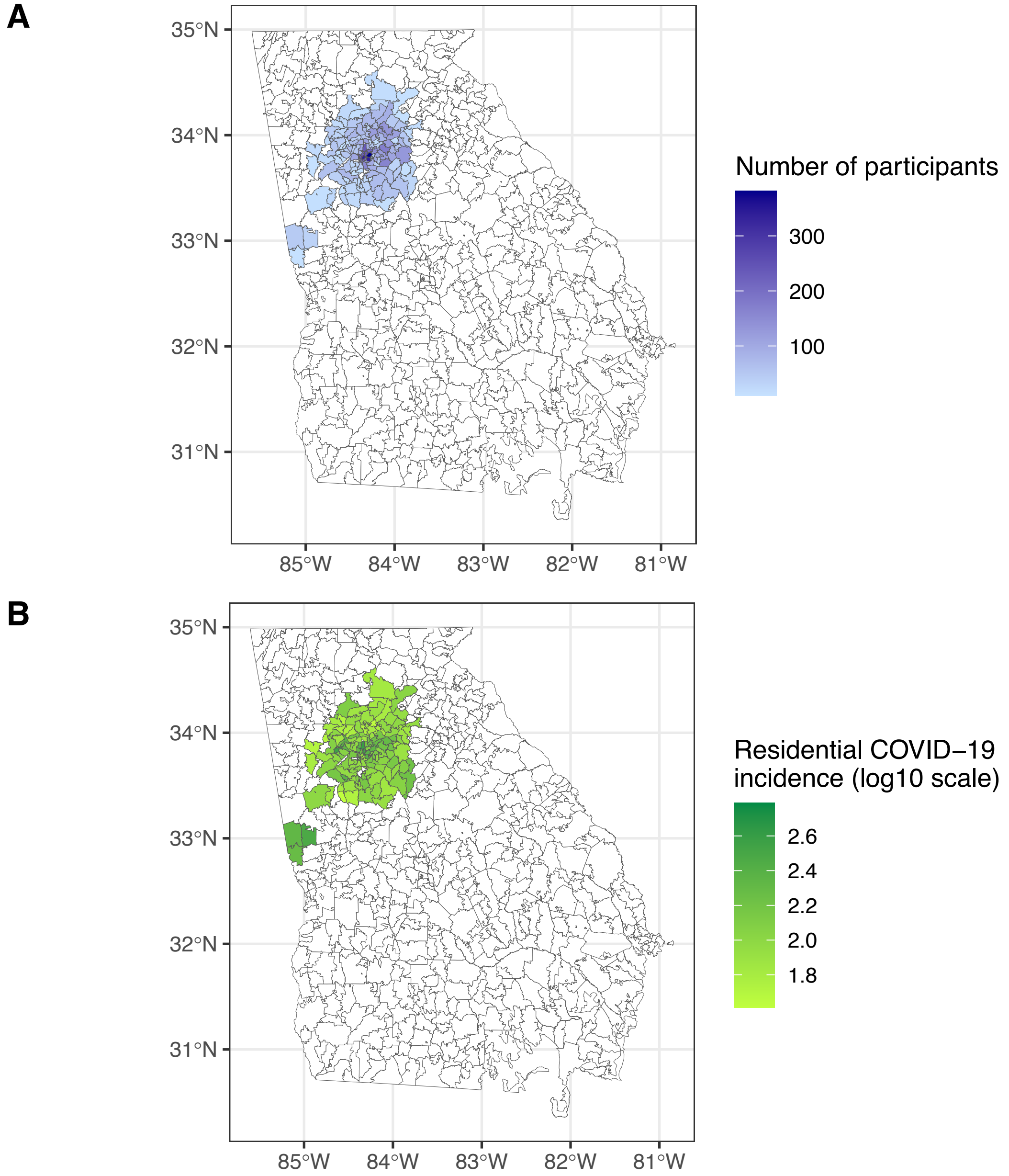


[a] Data for zip codes with fewer than 10 study participants are excluded from the figure.

**eTable 1. Details of data sources and factor descriptions.**

| Factor | Data source | Survey question | Possible responses | Additional information |
|---|---|---|---|---|
| **Demographic and community factors** | | | | |
| Age group | Employment records | NA | NA | Categorized into deciles |
| Race | Employment records | NA | NA | Self-reported |
| Ethnicity | Employment records | NA | NA | Self-reported |
| Contact with confirmed/suspected positive individual(s) | Survey | Since February 1, 2020, have you had close contact (>10 minutes of face to face) with someone with confirmed or suspected COVID-19 outside of work? | 1. Yes exposed to someone with COVID-19;<br>2. Yes;<br>3. No;<br>4. Not sure | Categorized into yes/no (not sure categorized as "no") |
| Residential COVID-19 incidence | Georgia Department of Public Health and employment records (residential zip code) | NA | NA | Cumulative incidence of persons under investigation for COVID-19 linked with zip code data and week of serology test. Cumulative incidence two weeks prior to week of serology test was used to account for lag between infection and testing. |
| **Occupational factors** | | | | |
| Caring for COVID-19 positive patient(s) | Survey | While at work, did/do you provide direct patient care for any patient(s) known (confirmed) to be positive for COVID-19, regardless of location in the facility? | Yes;<br>No;<br>Unknown | Unknown categorized as "no" |

**eTable 1. Details of data sources and factor descriptions (continued).**

| Factor | Data source | Survey question | Possible responses | Additional information |
|---|---|---|---|---|
| Caring patient(s) found to be COVID-19 positive while not on isolation precautions | Survey | While at work, did you care for patient(s) who, while not on isolation precautions, were tested and found to be COVID-19 positive? | Yes;<br>No;<br>Not sure | Not sure categorized as "no" |
| Contact with other healthcare worker found to be COVID-19 positive | Survey | While at work, did you have close contact (>10 minutes face to face) to other healthcare worker(s) who were found to have COVID-19 infection during the time period you were exposed to them? | Yes;<br>No;<br>Not sure | Not sure categorized as "no" |
| Workplace location | Survey | Where do/did you spend most time at work in the hospital during the COVID-19 pandemic? (may check more than one if you split your time equally) | 1. Emergency department;<br>2. Medical-surgical focused on COVID-19 patients;<br>3. Medical-surgical-not focused on COVID-19 patients;<br>4. ICU focused on COVID-19 patients;<br>5. ICU-not focused on COVID-19 patients<br>6. Clinic focused on COVID-19 patients;<br>7. Clinic not focused on COVID-19 patients;<br>8. OR or peri-operative area;<br>9. Other procedure area;<br>10. Other hospital area;<br>11. Non-clinical area of hospital/clinic<br>12. Not in hospital or clinic;<br>13. Work from home: worked from home | Hierarchy to classify participant into one category (even if multiple job locations were selected) in the order below. Order based on anticipated encounters with COVID-19 positive patients.<br>1. Emergency department: emergency department<br>2. COVID-19 focused: medical-surgical focused on COVID-19 patients; ICU focused on COVID-19 patients; clinic focused on COVID-19 patients<br>3. Inpatient not COVID-19 focused: medical-surgical-not focused on COVID-19 patients; ICU-not focused on COVID-19 patients<br>4. Outpatient clinical: clinic not focused on COVID-19 patients<br>5. OR/procedure: OR or peri-operative area; other procedure area<br>6. Other hospital area: other hospital area<br>7. No patient contact: non-clinical area of hospital/clinic; not in hospital or clinic<br>8. Work from home: worked from home<br>9. Not specified: no job location selected in survey |

**eTable 1. Details of data sources and factor descriptions (continued).**

| Factor | Data source | Survey question | Possible responses | Additional information |
|---|---|---|---|---|
| Workplace role | Survey | What is your job role? | 1. Advanced practice provider<br>2. Environmental services<br>3. Nurse<br>4. Nurse technician<br>5. Other direct care provider<br>6. Other with no patient contact<br>7. Physician<br>8. Radiology technician<br>9. Respiratory therapist | |
| PPE use | Survey | While at work, please assess your use of workplace recommended personal protective equipment (PPE) while caring for patients with suspected or known COVID-19. | 1. Had numerous situations where I was not using recommended PPE;<br>2. Wore PPE but unsure if I was always using properly;<br>3. Always used recommended PPE and paid close attention to donning and doffing | |

NA: not applicable; ICU: intensive care unit; OR: operating room; PPE: personal protective equipment

**eTable 2. Additional descriptive statistics for survey participants (n=9,610).**

| Factor | Total sample N. (%) | Seropositive N. (%) |
|---|---|---|
| **SARS-CoV-2 testing** | | |
| Week of serology testing | | |
| 4/19/2020 | 852 (8.9) | 40 (4.7) |
| 4/26/2020 | 1060 (11.0) | 40 (3.8) |
| 5/3/2020 | 874 (9.1) | 52 (5.9) |
| 5/10/2020 | 1954 (20.3) | 126 (6.4) |
| 5/17/2020 | 2270 (23.6) | 172 (7.6) |
| 5/24/2020 | 1721 (17.9) | 57 (3.3) |
| 5/31/2020 | 569 (5.9) | 52 (9.1) |
| 6/7/20 - 6/21/20 | 310 (3.2) | 16 (5.2) |
| **Prior illness (fever or respiratory symptoms) since February 1, 2020 & symptoms** | | |
| Prior illness, no COVID-19 test | | |
| No | 7969 (82.9) | 449 (5.6) |
| Yes | 1641 (17.1) | 106 (6.5) |
| Prior illness, negative COVID-19 test | | |
| No | 8882 (92.4) | 511 (5.8) |
| Yes | 728 (7.6) | 44 (6.0) |
| Previous positive COVID-19 test | | |
| No | 9477 (98.6) | 431 (4.5) |
| Yes | 133 (1.4) | 124 (93.2) |
| Previous illness (regardless of COVID-19 testing status) | | |
| No | 7191 (74.8) | 298 (4.1) |
| Yes | 2419 (25.2) | 257 (10.6) |
| Any symptoms | | |
| No | 129 (1.3) | 12 (9.3) |
| Yes | 2290 (23.8) | 245 (10.7) |
| Not applicable | 7191 (74.8) | 298 (4.1) |
| **Additional demographics & detailed job location** | | |
| Sex | | |
| Female | 7456 (77.6) | 428 (5.7) |
| Male | 2154 (22.4) | 127 (5.9) |

**eTable 2. Additional descriptive statistics for survey participants (n=9,610) (continued).**

| Factor | Total sample N. (%) | Seropositive N. (%) |
|---|---|---|
| Job location (dichotomous) | | |
| Medical-surgical- focused on COVID-19 patients | 1160 (12.1) | 83 (7.2) |
| Medical-surgical- not focused on COVID-19 patients | 2024 (21.1) | 120 (5.9) |
| ICU- focused on COVID-19 patients | 1320 (13.7) | 87 (6.6) |
| ICU- not focused on COVID-19 patients | 1445 (15.0) | 99 (6.9) |
| OR or peri-operative area | 877 (9.1) | 51 (5.8) |
| Other procedure area | 609 (6.3) | 40 (6.6) |
| Emergency department | 1007 (10.5) | 67 (6.7) |
| Other hospital area | 1716 (17.9) | 102 (5.9) |
| Clinic- focused on COVID-19 patients | 267 (2.8) | 16 (6.0) |
| Clinic- not focused on COVID-19 patients | 2190 (22.8) | 115 (5.3) |
| Non-clinical area of hospital or clinic | 1582 (16.5) | 84 (5.3) |
| Not in hospital or clinic | 239 (2.5) | 12 (5.0) |
| Work from home | 895 (9.3) | 42 (4.7) |

ICU: intensive care unit; OR: operating room

**eTable 3.** STROBE Statement—checklist of items that should be included in reports of observational studies

| | Item No | Recommendation | Page No |
|---|---|---|---|
| **Title and abstract** | 1 | (*a*) Indicate the study's design with a commonly used term in the title or the abstract | 1 |
| | | (*b*) Provide in the abstract an informative and balanced summary of what was done and what was found | 1 |
| **Introduction** | | | |
| Background/ rationale | 2 | Explain the scientific background and rationale for the investigation being reported | 2 |
| Objectives | 3 | State specific objectives, including any prespecified hypotheses | 2 |
| **Methods** | | | |
| Study design | 4 | Present key elements of study design early in the paper | 2 |
| Setting | 5 | Describe the setting, locations, and relevant dates, including periods of recruitment, exposure, follow-up, and data collection | 2 |
| Participants | 6 | (*a*) *Cohort study*—Give the eligibility criteria, and the sources and methods of selection of participants. Describe methods of follow-up<br>*Case-control study*—Give the eligibility criteria, and the sources and methods of case ascertainment and control selection. Give the rationale for the choice of cases and controls<br>*Cross-sectional study*—Give the eligibility criteria, and the sources and methods of selection of participants | 2 |
| | | (*b*) *Cohort study*—For matched studies, give matching criteria and number of exposed and unexposed<br>*Case-control study*—For matched studies, give matching criteria and the number of controls per case | n/a |
| Variables | 7 | Clearly define all outcomes, exposures, predictors, potential confounders, and effect modifiers. Give diagnostic criteria, if applicable | 2-3 |
| Data sources/ measurement | 8* | For each variable of interest, give sources of data and details of methods of assessment (measurement). Describe comparability of assessment methods if there is more than one group | 2-3, eTable 1 |
| Bias | 9 | Describe any efforts to address potential sources of bias | 3 |
| Study size | 10 | Explain how the study size was arrived at | 2 |
| Quantitative variables | 11 | Explain how quantitative variables were handled in the analyses. If applicable, describe which groupings were chosen and why | 2-3, eTable 1 |
| Statistical methods | 12 | (*a*) Describe all statistical methods, including those used to control for confounding | 2-3 |
| | | (*b*) Describe any methods used to examine subgroups and interactions | n/a |
| | | (*c*) Explain how missing data were addressed | 3 |
| | | (*d*) *Cohort study*—If applicable, explain how loss to follow-up was addressed<br>*Case-control study*—If applicable, explain how matching of cases and controls was addressed<br>*Cross-sectional study*—If applicable, describe analytical methods taking account of sampling strategy | n/a |
| | | (*e*) Describe any sensitivity analyses | 2-3 |

**eTable 3.** STROBE Statement—checklist of items that should be included in reports of observational studies (continued)

| | | | |
|---|---|---|---|
| **Results** | | | |
| Participants | 13* | (a) Report numbers of individuals at each stage of study—eg numbers potentially eligible, examined for eligibility, confirmed eligible, included in the study, completing follow-up, and analysed | 3 |
| | | (b) Give reasons for non-participation at each stage | 3 |
| | | (c) Consider use of a flow diagram | n/a |
| Descriptive data | 14* | (a) Give characteristics of study participants (eg demographic, clinical, social) and information on exposures and potential confounders | 3, Table 1, eTable 2 |
| | | (b) Indicate number of participants with missing data for each variable of interest | Table 1 |
| | | (c) *Cohort study*—Summarise follow-up time (eg, average and total amount) | n/a |
| Outcome data | 15* | *Cohort study*—Report numbers of outcome events or summary measures over time | n/a |
| | | *Case-control study*—Report numbers in each exposure category, or summary measures of exposure | n/a |
| | | *Cross-sectional study*—Report numbers of outcome events or summary measures | 3, Table 1 |
| Main results | 16 | (*a*) Give unadjusted estimates and, if applicable, confounder-adjusted estimates and their precision (eg, 95% confidence interval). Make clear which confounders were adjusted for and why they were included | 3-4, Table 1 |
| | | (*b*) Report category boundaries when continuous variables were categorized | n/a |
| | | (*c*) If relevant, consider translating estimates of relative risk into absolute risk for a meaningful time period | n/a |
| Other analyses | 17 | Report other analyses done—eg analyses of subgroups and interactions, and sensitivity analyses | 3-4 |
| **Discussion** | | | |
| Key results | 18 | Summarise key results with reference to study objectives | 4-5 |
| Limitations | 19 | Discuss limitations of the study, taking into account sources of potential bias or imprecision. Discuss both direction and magnitude of any potential bias | 5 |
| Interpretation | 20 | Give a cautious overall interpretation of results considering objectives, limitations, multiplicity of analyses, results from similar studies, and other relevant evidence | 4-5 |
| Generalisability | 21 | Discuss the generalisability (external validity) of the study results | 4-5 |
| **Other information** | | | |
| Funding | 22 | Give the source of funding and the role of the funders for the present study and, if applicable, for the original study on which the present article is based | 5 |

*Give information separately for cases and controls in case-control studies and, if applicable, for exposed and unexposed groups in cohort and cross-sectional studies.
